# Frequency of testing for COVID 19 infection and the presence of higher number of available beds per country predict outcomes with the infection, not GDP of the country – A descriptive statistical analysis

**DOI:** 10.1101/2020.04.01.20047373

**Authors:** Binayak Sinha, Sumit Sengupta, Samit Ghosal

**Author notes:** Corresponding author: Samit Ghosal.

## Abstract

**Introduction:** The novel coronavirus epidemic which originated in late 2019 from China has wreaked havoc on millions across the world with illness, death and socioeconomic recession. As of now no valid treatment or preventative strategy has evolved worldwide and governments across the world have been forced to take the draconian step of social isolation in communities by enforcing “lockdowns”.

**Aim of this Study:** This study aims to correlate the rates of infection with the novel coronavirus and total deaths as the primary output variable. In addition the strength of association between infection rates and total death in comparison to GDP share of the respective countries, physicians, hospital beds and rates of testing for COVID 19 infection per thousand patients, is being assessed, in a bid to develop a model which would help to develop tools to reduce the impact of this disease.

**Material & Methods:** Data relating to number of cases, severity, cases recovered and deaths worldwide and specifically for the top six countries affected was collected from the WHO COVID-19 situation report which is being updated on a daily basis till 22^nd^ March 2020, the date of analysis. Additional data related to GDP, physician and hospital bed per 1000 patients were procured from the World Bank database. All data were collected in a file in CSV format. Analysis was conducted in Jupyter notebook with Python 3.8.2 software and also with XL-Stat statistical software for excel. The analytical strategy was descriptive with no inferential overtones.

**Results:** COVID 19 infection strongly correlates with total deaths (r : 0.89), with a predicted death rate of 25 patients per 1000 affected. There was no correlation between the GDP growth of the country and number of treating physicians/1000 patient population with any COVID 19 related outcome. However there was a negative correlation between COVID 19-related deaths and the number of beds available per 1000 population [r=-0.34]. Importantly there is an inverse correlation between the number of tests conducted per million population with the rates of active infections [r=-0.12], new cases [r=-0.38] and new deaths [r=-0.28] in COVID 19.

**Conclusion:** This is the first study to assess parameters other than age and sex and sets out a robust dataset which indicates an increased risk of worsening outcomes with lesser number of beds and testing, suggesting that the need of the hour is to increase available bed numbers and to increase rates of testing.

## 1. Introduction

The outbreak of coronavirus pandemic which started from late 2019 has created a havoc on human civilization. We are not only faced with the disease-related morbidity and mortality, but also extensive economic, social and psychological upheaval. In view of the rapidity of spread of infection and a high mortality rate the present day health-care system is firmly focussed on both prevention and treatment. Research required to arrive at a definitive therapeutic intervention is going to take some time, but there are encouraging reports from non-randomised trials with drugs like chloroquine/hydroxychloroquine, remdesivir, azithromycin.[1,2] Governments across the globe have taken on this calamity of herculean proportions head on with lockdown of cities in an attempt to contain further spread of the disease.

However, it is also extremely important to analyse the evolving data at hand to get an understanding of the direction we are headed. Such an analysis might also help us prepare more effectively to plan for the future and fortify preventive strategies giving researchers ample space and time to come up with potent and effective cure for COVID-19 pneumonia.

## 2. Aims of this analysis

This analysis aims at finding a correlation between the rates of infection and total death. In addition we also looked at the strength of association between infection rates and death in comparison to GDP share of the respective countries, physicians & hospital beds/1000 patients and rates of testing for COVID 19 infection per 1 million patients. Lastly, we plan to create a regression model which would help predicting the death rates in those infected with COVID-19.

## 3. Materials & Methods

We collected the data from the WHO COVID-19 situation report which is being updated on a daily basis.[3] Since we needed to do the analysis at a point in time we selected the data available as of 22^nd^ March as the date of analysis. Since data relating to age and sex and their correlation to COVID 19 outcomes have already been published, those data were not collated [4]. Additional data related to GDP, physician and hospital bed per 1000 patients were procured from the World Bank database. [5,6,7]

All data were collected in a file in CSV file format. Analysis was conducted in Jupyter notebook with Python 3.8.2 software and also with XL-Stat statistical software for excel.

The analytical strategy was descriptive with no inferential overtones.

## 4. Results

### 4.1 Patients infected with COVID-19 and mortality

A very strong correlation (r: 0.89) was found between the patients infected with COVID-19 and total death. (Fig 1) There was a 89% association between death and getting infected (P<0.001, 95% CI: 0.046-0.056).

**Fig 1:**
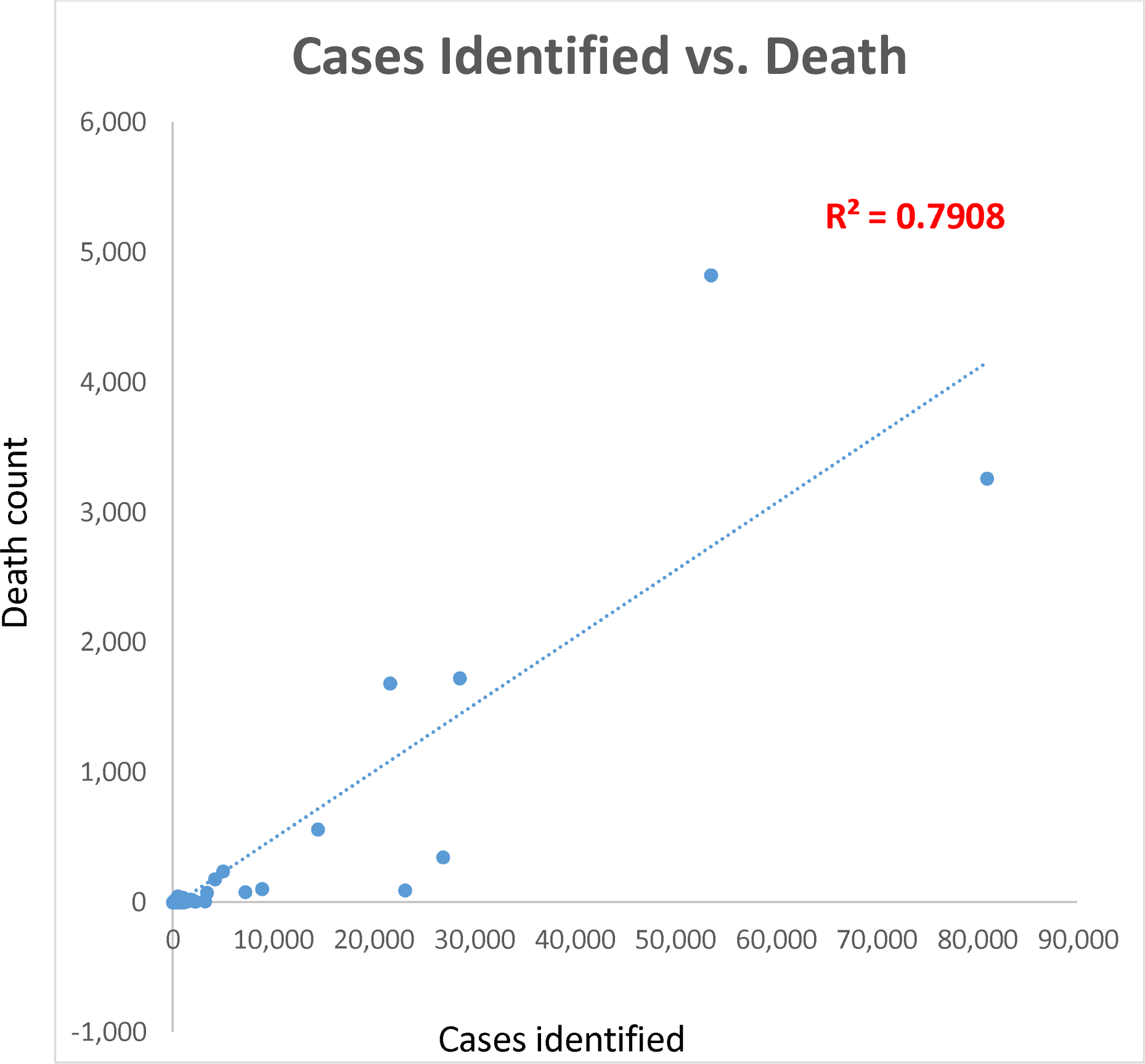
Scatter plot comparing Death and Cases identified (COVID-19) with correlation data: Linear regression analysis with line of best fit.

For every 1000 patient the predictive death count was 25.

The regression model for prediction was: y(total number of death from COVID-19) = -26.72 + 0.05*total infected population. (Fig 2)

**Fig 2:** Regression analysis with regression model comparing total death and those infected with COVID-19: Raw data.

### 4.2 Relationship between GDP, physician and hospital bed per 1000 patient population and infection rate & mortality

There was poor correlation between GDP growth and infection rate [r=0.04] or death from COVID-19 [r=0.01]. As a matter of fact the top 5 infected countries bearing most of the brunt from COVID-19 infection shares 48% of the Global GDP between them. (Table 1)

**Table 1:** Comparison of total cases and GDP & Physician and hospital bed per 1000 patient population from the top 6 infected countries.

| *Country* | Total Cases | Share of World GDP | Hospital beds per 1000 patients | Physicians per 1000 patients |
| --- | --- | --- | --- | --- |
| *China* | 81,054 | 0.1512 | 3.56 | 1.7855 |
| *Italy* | 53,578 | 0.024 | 3.6 | 3.9174 |
| *Spain* | 28,572 | 0.0162 | 3.2 | 3.8493 |
| *United States* | 26,909 | 0.2408 | 3 | 2.5858 |
| *Germany* | 23,129 | 0.0456 | 8.2 | 4.1383 |

In view of the wide heterogeneity of data entry across the different countries including most of the input parameters there were no correlation trend visible as far as infection rates and its relationship with the physicians or hospital bed per 1000 patient population was concerned. (Fig 3) However, analysing the top 15 infected countries gave a definitive trend. (Fig 3) New deaths from COVID 19 was inversely correlated with the number of available hospital beds per 1000 infected patients admitted [r=-0.34]. However, there was no trend observed between the COVID-19 related deaths and physicians per 1000 population[.r=-.011]. (Fig 3)

**Fig 3:** Correlation statistics from the top 15 infected countries with testing frequency included.

Amongst the top 6 infected countries Germany had the best physician and hospital bed per 1000 patient population.

### 4.3 Correlation between testing for COVID-19 and other variables

In view of increased frequency of testing seen predominantly in the top 15 countries, analysis was done on the same to improve the yield of outcomes. Increased testing frequency was associated with a positive correlation with recovery [r=0.18] and was negatively correlated with new cases, [r=- 0.38] new deaths, [r=-0.28] active cases [r=-0.12]. (Fig 3)

## 5 Discussion

The number of people diagnosed with COVID-19 have crossed 3 lakhs with more than 12,000 deaths at the time of this analysis. The figures keep climbing up steeply with each passing days. It is a crisis of mammoth proportion requiring the cooperation from the individual to the global leaders in health and politics. The situation came into focus when 44 cases of pneumonia of unknown cause were reported from the Wuhan province of China. [8] It was the beginning of the second week of January when a novel strain of coronavirus was found to be responsible. Thereafter there has been a steep rise in cases not only in the Hubei province but different regions of China and adjoining countries like Thailand. With increasing spread of the disease and the associated number of deaths, different countries started to react in an attempt to prevent the disease. The approaches included case finding, isolation of suspected cases and their close contacts and also hunting for active therapeutic agents to tackle the viral-related morbidity and mortality.

Recent genomic studies have practically ruled out the possibility of a laboratory related experiment gone out of hand. [9] The two hypothesis doing rounds are a non-pathogenic strain of coronavirus which jumped into a human host from an animal intermediary and became pathogenic by the process of natural selection and the other possibility being a virus becoming pathogenic by the process of natural selection in an animal species and then jumps on to its human host [9]. Having identified the genomic sequence and its predilection for the lungs, the next step was to find a diagnostic test and discover medications to tackle the situation. [10]

However, a more important aspect of the disease in question has to do with prevention. Identifying the appropriate factors related to getting infected and mortality would help plan in advance. Till now the only valid and detailed analysis and correlation data relating to COVID 19 outcomes has been related to age and sex variables. [11]

This is the first analysis to ascertain certain additional factors related to adverse outcomes associated with COVID-19 infection. What is the correlation and its strength as far as getting infected and mortality were concerned? Can we build a predictive model on the same using a linear regression analysis? What is the recovery rate country-wise? Is there a difference depending on the geographical region? How does GDP share of the country in question and the number of physicians & hospital bed per 1000 patient available impacts survival? These are crucial questions which could help us identify important logistics related factors contributing to the direction the infected population would go.

We found a statistically significant 89% association with death once an individual gets infected with COVID-19. There were 25 expected deaths per 1000 patient infected. This is in keeping with the case fatality rates that are evolving worldwide, [12] with a significant percentage of these deaths effecting the elderly.[13] With the help of regression analysis we could build a mathematical model to predict death depending on the number infected.

Germany has a very low death rate in spite of being on the top 6 countries being affected. This could be attributed to the highest physician and hospital bed per 1000 patient ratio among the top 6 countries. It should also be noted here that recovery rates in China improved dramatically after an increase in the number of beds made available, along with strong quarantine measures that were applied. This analysis bolsters the notion that higher number of hospital beds per 1000 population corelates with a lower death rate. This might provide the basis to make an appropriate calculation and get a proportional amount of beds prepared in advance.

It is interesting to note that GDP share has no correlation whatsoever with both the infection rates as well as mortality. Wealth and economic resources may therefore not be adequate in tackling the scourge of this novel coronavirus.

What is most revealing about this data, however, is that higher testing rates improved recovery rates. Increased number of tests were significantly correlated with 1) a reduction in the number of new cases, 2) a reduction in the number of active cases and 3) most importantly with a reduction in the number of critically ill patients. This would indicate that the best option to deter this pandemic is higher number of tests being conducted in all populations. Intuitively one could surmise that increasing tests would identify the infected individuals early in the disease process providing more robust and aggressive treatment to this population, improving morbidity and probably mortality.in addition it would warn the exposed to take strict steps to quarantine themselves, reducing rates of new and active cases.

### 5.1 Study limitations

Firstly, correlation studies do not mean causality. The associations mentioned above are indicative of certain trends only. However, at an early stage of a novel disease trends can definitely be a good approximation for a large data-related analysis.

Secondly, due to extreme paucity of data from many countries, they were excluded in certain analysis for example the one on testing frequency and outcomes. This weakness was overcome by including the top 15 countries wherefrom the majority of the data were more homogenous and devoid of significant outliers.

Thirdly, the correlation values are not indicative strong associations-both positive and negative. However, this is due to the fluidity of the situation. We do not desire the numbers to go up to give us opportunity for a robust analysis. This is precisely why we attempted to identify trends from the updated data and guide our health care system to gear up for a better assessment of the situation.

Fourthly, the use of GDP as a measure of disease outcomes cuts both ways. If it is an indicator of surplus wealth, then it is expected that a large proportion of the same could be channelized into the health care facility and hence the outcomes. Hence, we choose to include both GDP as well as share of the World’s GDP in this analysis to overcoming this confounding.

### 5.2 Strength of the study

In contrast to the association data related to mortality rates and risk factors for the same like sex age, other co-morbidities, this analysis is probably the first in its kind to look at the correlation between infection rate/mortality and GDP, physician & hospital bed per 1000 patient population, testing frequency and its relation with reduced new cases being detected as well as new deaths being prevented. Although these are only trends, we believe these data will open up newer avenues for health-care sector planning leading to effective management strategies.

## 6. Conclusion

This is the first study to indicate that increasing resources in the form of increasing number of beds per 1000 population reduces death from COVID 19 and improves recovery thereof. More importantly this is the first dataset analysis which reveals that increasing number of tests for the novel coronavirus would drastically reduce incidence and possibly death rates and morbidity from this dreaded epidemic.

## Data Availability

All data are available in CSV file format and can be provided on request.

## Acknowledgements

- Mr. Milan Majumder: Independent statistician, Pune, India
- Mr. Kingshuk Bhattacharjee: Independent Biostatistician, Kolkata, India.

## Figure & Table Legends

### SUMMARY

#### OUTPUT

| Regression Statistics |  |
| --- | --- |
| Multiple R | 0.889290998 |
| R Square | 0.79083848 |
| Adjusted R Square | 0.788704178 |
| Standard Error | 286.4700615 |
| Observations | 100 |

#### ANOVA

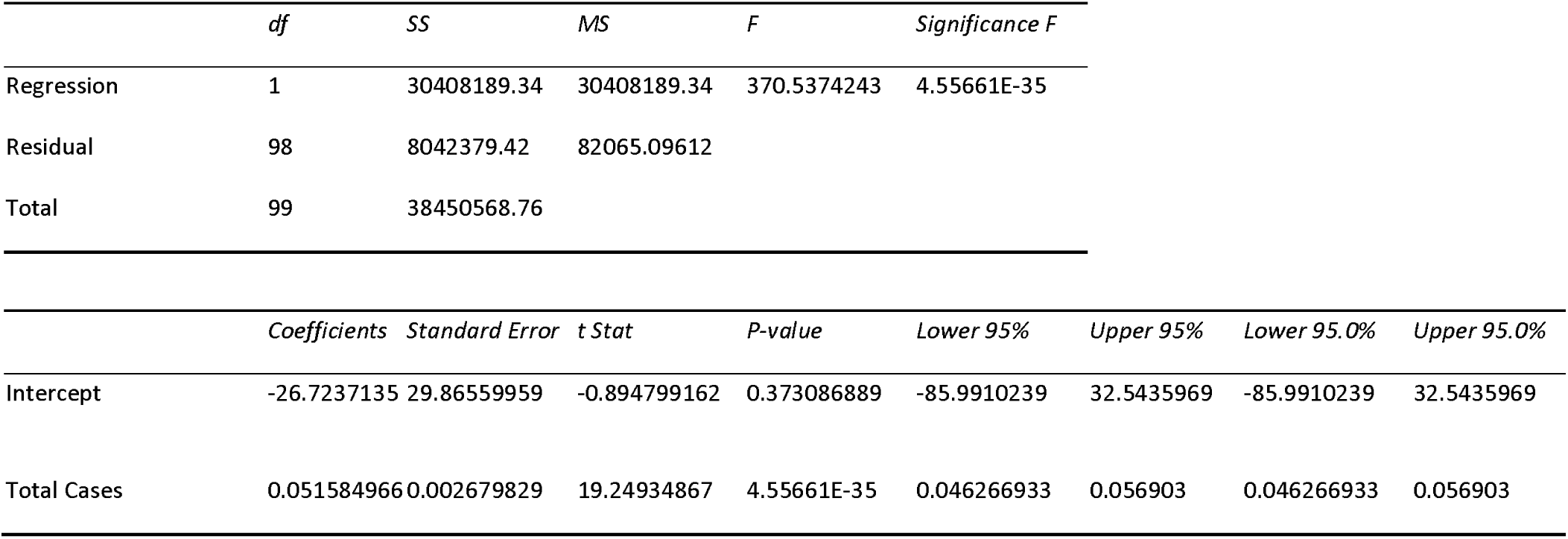

### SUMMARY

#### OUTPUT

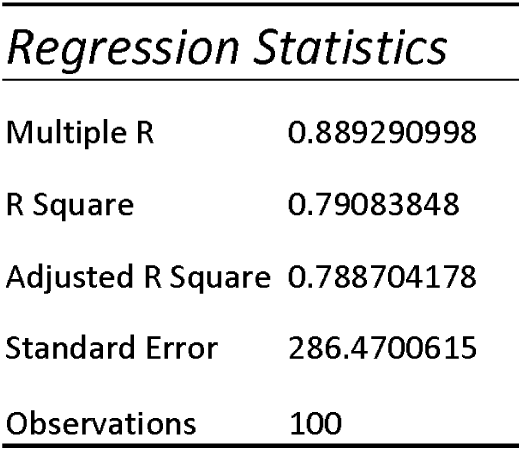

SUMMARY OUTPUT
| *Regression Statistics* |  |
| --- | --- |
| Multiple R | 0.889290998 |
| R Square | 0.79083848 |
| Adjusted R Square | 0.788704178 |
| Standard Error | 286.4700615 |
| Observations | 100 |

#### ANOVA

|  | *df* | *SS* | *MS* | *F* | *Significance F* |
| --- | --- | --- | --- | --- | --- |
| Regression | 1 | 30408189.34 | 30408189.34 | 370.5374243 | 4.55661E-35 |
| Residual | 98 | 8042379.42 | 82065.09612 |  |  |
| Total | 99 | 38450568.76 |  |  |  |

|  | *Coefficients* | *Standard Error* | *t Stat* | *P-value* | *Lower 95%* | *Upper 95%* | *Lower 95.0%* | *Upper 95.0%* |
| --- | --- | --- | --- | --- | --- | --- | --- | --- |
| Intercept | -26.7237135 | 29.86559959 | -0.894799162 | 0.373086889 | -85.9910239 | 32.5435969 | -85.9910239 | 32.5435969 |
| Total Cases | 0.051584966 | 0.002679829 | 19.24934867 | 4.55661E-35 | 0.046266933 | 0.056903 | 0.046266933 | 0.056903 |

